# Cell-wall-deficient Bacteria in Oral Biofilm: Association with Periodontitis

**DOI:** 10.1101/2020.07.13.20120428

**Authors:** Francesco Germano, Davide Testi, Luisa Campagnolo, Manuel Scimeca, Claudio Arcuri

## Abstract

Cell-wall-deficient bacteria are those that lack cell walls and live in a pleomorphic state. The genus Mycoplasma and L-form bacteria are both members of this group. The aim of this study was to search cell-wall-deficient bacteria in periodontal biofilm and link their presence to periodontal disease. Eighty-nine individuals were recruited and divided into three groups: periodontally healthy individuals, individuals with chronic periodontitis, and those with aggressive periodontitis. The presence of cell-wall-deficient bacteria was detected in freshly collected biofilm by light microscopy, transmission electron microscopy (TEM) with and without electron microscopy in situ hybridization, atomic force microscopy and DNA stain (Hoechst). A new dichotomic index of classification for prevalence and morphologic variants was developed to classify cell-wall-deficient bacteria in periodontal biofilm. Cell-wall-deficient bacteria were found in periodontal biofilm and classified into Protoplastic, Everted, Filament and Intracellular forms, the last one mainly associated with aggressive periodontitis. We also assessed the prevalence of periodontopathic bacteria by means of polymerase chain-reaction (PCR) and found no clear, statistically significant, correlation among periodontal pathogens tested (except *T. denticola*) that allowed individuals with chronic periodontitis to be distinguished from those with aggressive periodontitis. Association between cell-wall-deficient bacteria and periodontal condition was: periodontally healthy, 3.3% (1/30); individuals with chronic periodontitis, 30.6% (11/36); and those with aggressive periodontitis, 100% (23/23). Cell-wall-deficient bacteria were detected in periodontal biofilm and linked to aggressive periodontitis.

## Introduction

Since the middle of the last century, there has been a rapid knowledge evolution in the field of microbiology that is reflected in a non-homogeneous literature. Analyzing mid-1900s studies on “non-classical” bacterial forms, we found terms such as ‘cell-wall-deficient bacteria’ (CWDB), ‘pleuropneumonia-like organisms (PPLO)’, ‘mycoplasma’, ‘L-form bacteria’, ‘spheroplast’, and ‘protoplast’ used to describe pleomorphic forms of different bacteria.

Since the discovery of CWDB, several studies have linked their presence to different diseases, but their role remains unclear. Some authors believe that the ability of bacteria to lose the cell wall and to replicate in the CWD state represents a primordial reproduction mechanism [[1]]. We now know that the bacteria that can colonize the oral cavity belong to more than 700 species [2]. Some researchers have stated that probably all known bacterial species can be converted to CWDB by a variety of inducing agents such as cell-wall-inhibiting antibiotics and high concentrations of amino acids and enzymes [3-4]. No studies are reported on oral CWDB.

Cell-wall-deficient bacteria (CWDB) are those that lack a cell wall, live in a pleomorphic state, and proliferate by a mechanism of membrane tubulation, blebbing, or vesiculation [1,3]. The genus Mycoplasma and L-form bacteria are both members of this group. ‘Mycoplasma’ is the term now used to describe a heterogenic group of bacteria (members of mollicutes), of very small genome size, that have evolved from walled bacteria (members of firmicutes). The clinical picture of mycoplasma infections in humans and animals is more suggestive of damage due to host immune and inflammatory responses rather than to direct toxic effects by mycoplasmal cell components [5].

*Mycoplasma salivarium* (or the pleuropneumonia-like organism PPLO) was found in the oral cavities of all individuals with natural dentition and was absent in edentulous individuals [6]. *M. salivarium* can play an etiologic role in some oral infections, including periodontal diseases [7]. One study reported that the proportion of mycoplasmas is related to periodontal disease severity [8]. Mycoplasmas have been detected in a patient diagnosed as having juvenile periodontitis [9]. L-form bacteria were described for the first time more than 80 years ago [10] and can be differentiated into stable or unstable, depending on their ability to revert to the parental, cell-walled form [11]. Recent reports hypothesized that early key steps in the evolutionary development of the bacterial domain of life have been achieved by CWD bacteria, and that the mechanism whereby bacteria can live without a cell wall may have been retained by modern cells as a back-up process for use when cell wall synthesis is compromised [1,12]. Reversible model of transient cell wall-deficiency based on E. coli that allows studying transition, L-form reproduction and reversion in a controlled manner has been recently presented [13]. Some authors described the survival of *E. coli* L-forms under lethal stress, whether by autoclaving or boiling [14]. Since their identification, numerous reports have linked CWD bacteria with the persistence and recurrence of many diseases such as osteomyelitis, rheumatism, endocarditis, Alzheimer’s disease, Lyme disease, and even cancer [4,15-18]. Ultrastructure analysis has been used to identify Mycoplasma [19] and L-forms of bacteria [20] in cell culture.

Periodontitis is a disease caused by lack of homeostasis between complex mixed-species periodontal biofilm and host multifactorial response. Periodontal disease manifestation varies from mild chronic to severe aggressive, depending on still-not-completely-understood predisposing factors. In a systematic review [21], the authors found no clear correlation among periodontal pathogens that allow individuals with chronic periodontitis (CP) to be distinguished from those with aggressive periodontitis (AP). In another study, the correlation among periodontal pathogens resulted in large differences in the composition of subgingival plaque between CP and AP [22].

Since literature associating CWD bacteria with periodontal disease is almost non-existent, we have attempted to verify the hypothesis that in chronic and aggressive forms of the disease, the relationship between periodontal biofilm and inflammatory host response is linked to the presence of CWD bacteria. The purpose of this study was to classify the different forms of CWDB present in periodontal biofilm and associate their presence with periodontal disease. We first assessed the prevalence of major periodontopathic bacteria, in our samples, by means of polymerase chain-reaction (PCR). We then developed a simple method to identify and classify CWD bacteria in freshly collected biofilm using phase-contrast microscopy and evaluated distribution of DNA in these microorganisms using Hoechst dye. The presence of CWDB in periodontal biofilm was also assessed after dissociated-biofilm culture.

In a previous study [23], we used Atomic Force Microscopy (AFM) on oral biofilm to study morphological characteristics of some periodontal bacteria at high magnification with simple and conservative sample preparation. In this study, we used AFM to identify morphological characteristics of CWDB. Finally we confirmed the presence of CWDB in periodontal biofilm by transmission electron microscopy (TEM) with and without in situ hybridization,

## Materials & Methods

### Study Population

Participants were diagnosed and then categorized into 3 clinical groups according to AAP periodontal classifications [22]. We set an age cut off point of 35 years and verify the presence of two or more interproximal, nonadjacent sites with attachment loss of ≥ 6 mm occurring at a minimum of two different teeth and accompanied by bleeding on probing for aggressive periodontitis as suggested in recent reviews [24-25]. Eighty-nine individuals were randomly selected from among those referred to the periodontal and general dentistry department at the Fatebenefratelli “S. Giovanni Calibita” hospital in Rome, Italy, and were divided into groups as follows: 30 periodontally healthy (mean age, 51 yrs; age range, 25-68 yrs; males, 15; females, 15), 36 with chronic periodontitis (mean age, 57.2 yrs; age range, 36-75 yrs; males, 22; females, 14), and 23 with aggressive periodontitis (mean age, 31,8 yrs; age range, 24-35 yrs; males, 6; females, 17).

For each patient, the following data were recorded and used for classification: bleeding index, plaque index, probing depth, and clinical attachment level. Periodontal examination was carried out by a single trained and calibrated examiner (TD). Calibration of clinical periodontal parameters was carried out on five participants (k value 0.83). Individual data (PD, CAL, PI, BOP, etc.) for all patients are presented in the online Supporting Information (Table S1).

Individuals were excluded if: they had fewer than 16 teeth, they had taken antibiotics in the preceding 3 mos, they had received periodontal treatment in the preceding 6 mos, they were smokers, or they had systemic conditions affecting their periodontal status according to AAP classifications.

Written informed consent was obtained from each patient. Examinations on patients and samples of Oral Biofilm were part of routine and diagnostic care and used for statistical purpose. All data was anonymized prior to analysis.

The Review Board of the Ethics Committee at the Fatebenefratelli “S. Giovanni Calibita” hospital, in charge of preliminary research activity, was informed of the study, examined the documents, approved this consent procedure and provide an oral waiver for the proposed protocol. However the Review Board decided to not submit the proposed protocol to the Ethics Committee for a formal approval because sample and analysis on Oral Bacteria is considered normal clinical practice, it was believed that the issues did not meet those expressly covered by the rules.

### Microbiological Samples

Biofilm samples were collected, by means of a sterile curette, from the deepest pocket area of individuals with periodontitis after careful removal of supragingival plaque. Samples from individuals with periodontitis were pooled samples collected from the 4 deepest pockets, one *per* quadrant, and samples from periodontally healthy individuals were pooled samples collected from 4 first molars (or second molars if the first was missing), one *per* quadrant. The biofilms were dispersed in 200 μL of sterile saline solution (0.9%) by means of a sterile hypodermic syringe, and a 50 μL quantity of the pool obtained was applied to a glass slide and covered with a coverslip. Excess solution was removed from the edge of the coverslip, and the slide was sealed with nail varnish. Microbiological samples were analyzed by two operators (GF, TD) blinded to the clinical status of the study participants. Both operators were trained to look for CWDB according to our classification, and frequency of agreement was 93% (k value 0.87). A microscopic image was obtained with an optical microscope in phase contrast at 200x, 400x, and 600x magnifications (BA310 LED Motic Asia, Hong Kong). The images and movies were captured with a digital camera (Canon 7D). Two biofilm samples for each clinical group were also dispersed in Luria-Bertani broth (LB) containing 40 mM MgCl2, 1 M sucrose and 40 mM maleic acid pH 7 to test whether a higher level of osmotic protection allows better preservation of CWDB. Seven biofilm samples (2 from periodontally healthy individuals, 2 from chronic periodontitis individuals, 3 from aggressive periodontitis individuals) were processed for bacterial culture. Nine biofilm samples (3 from periodontally healty individuals, 3 from chronic periodontitis individuals, 3 from aggressive periodontitis individuals) were processed for transmission electron microscopy (TEM) and electron microscopy in situ hybridization.

### PCR

Samples for PCR were collected from individuals with chronic and aggressive periodontitis and processed as previously described [26]. The amplified DNA was applied to the DNA-Strip^®^ matrix to test the prevalence of periodontopathic bacteria (micro-IDent ® plus for *A. actinomycetemcomitans, P. gingivalis, P. intermedia, T. forsythia, T. denticola F. nucleatum, C. rectus, P. micra, E. nodatum, E. corrodens*, and *Capnocytophaga* sp.; Hain Lifescience GmbH, Nehren, Germany)

### TEM

Biofilm samples collected as reported above were fixed in Karnovsky’s solution (2% paraformaldehyde / 2% glutaraldehyde) for 24h, embedded in 2% agarose and post-fixed in 2% osmium tetroxide. After washing with 0.1 M phosphate buffer, samples were dehydrated by a series of incubations in 30%, 50% and 70%, ethanol. Dehydration was continued by incubation steps in 95% ethanol, absolute ethanol and propylene oxide, then samples were embedded in Epon (Agar Scientific, Stansted Essex CM24 8GF United Kingdom) [27]. After incubation at 60°C for 48h, epon embedded cells were cut by diamond knife (DiaTome, 1560 Industry, Hatfield, PA 19440).

In detail, 60-80 nm ultrathin sections (silver-yellow color) were collected onto cupper grids and staining with heavy metals solutions as previously described [28].

All samples were observed by TEM Hitachi FA 7100

### Pre-embedded electron microscopy *In Situ* Hybridization

After fixation in Karnovsky’s solution small fragment of samples were treated for *In Situ* Hybridization. Briefly, for bacterial permeabilization, samples were incubated with both lysozyme solution (Thermo Scientific, Waltham, MA USA) and Triton X-100 (Thermo Scientific, Waltham, MA USA) at room temperature for 1h. Then, samples were pre-treated with EDTA citrate Ph 7.8 (UCS Diagnostics, Rome, Italy) at 95°C for 20 min, and incubated with EUB 338 biotinylated probe (5’ GCT GCC TCC CGT AGG AGT 3’) (Eurofins Genomics, Ebersberg, Germany) for the detection of Bacterial 16S rRNA in a humid chamber for 24h at 37 °C. Once completed hybridization step, samples were washed in 50 ml of stringent washing buffer (Abbot, Illinois, U.S.A) at 37 °C for 30 min. Whereupon, samples were incubated with streptavidin-gold nanoparticles (20nm) conjugated (Agar Scientific, Stansted Essex CM24 8GF United Kingdom) for 2h at room temperature. Finally, all samples were embedded in Epon and observed by TEM as described above.

### Atomic force microscopy

Biofilm samples for atomic force microscopy were collected as previously described [23]. Images were taken with AFM SolverPro-M^®^, equipped with ETALON HA_NC/15 probes (NT-MDT, Moscow, Russia) in semi-contact and phase modes, and scanned *per* sample. All images were acquired at 1,024 lines. Image analysis was performed with the free software Gwyddion^®^ (http://gwyddion.net).

### Fluorescence Staining

Hoechst 33342 (0.5 μg mL^-1^; Sigma-Aldrich, St. Louis, MO, USA) was added to 50 μL of six biofilm samples pool obtained from individuals with chronic periodontitis, aggressive periodontitis and from periodontally healty individuals. Slides were mounted with Mowiol^®^, and fluorescent images were taken with a Zeiss Axioplan2 microscope.

### Bacterial Culture

Biofilm samples collected as reported above were transferred in a 15 ml sterile tube containing 5 ml of Luria-Bertani broth (LB) and incubated for 2 days at 30°C without shaking. Four millilitres of each culture were filtered using a sterile 0.45 μm filter (Starstedt, Germany) to exclude walled bacteria and eukaryotic cells. To concentrate samples, filtered material was centrifuged and resuspended in 50 μl of LB to which 1 μl of Hoechst (stock solution 1 mg/ml) was added. Samples were spotted on a slide for microscopy analysis. Phase contrast and fluorescent images were taken with a Zeiss Axioplan2 microscope connected to a digital camera.

### Index for CWDB Classification

The nature of CWD L-form bacteria, with no homogeneous distribution inside the biofilm, and their high polymorphism (intra- or extracellular and a large size range) prevented us from using traditional counting methods; therefore, we created a CWDB classification index based on descriptions present in the literature [29]. We identified 4 different morphological varieties of CWDB: protoplastic with or without inclusion, everted, filament, and intracellular forms.

Based on the quantity and quality of CWDB present, we developed a dichotomic index to assess their prevalence in periodontal biofilm:

*Grade 0* (G0) was assigned when, in 10 random fields (100 x 100 microns), the absence of CWDB or the presence of only protoplastic form without granules < 1% of total bacteria was recorded; and

*Grade 1* (divided into high and low concentrations, G1L, G1H) was assigned when, in 10 random fields (100 x 100 microns), one of the following conditions was met:

- G1L - when 2 kinds of CWDB were present, or one or more areas of the slide had CWDB prevalence > 5%; or
- G1H - when 3 or more kinds of CWDB were present, or one or more areas with high prevalence of CWDB were found (> 70%).

### Statistical Analysis

Differences in the prevalence of CWDB in healthy individuals and those with chronic or aggressive periodontitis were analyzed by the chi-square test with Prism 6 GraphPad software. Cohen’s kappa coefficient was used for calibration of clinical periodontal parameters and for index score agreement between operators.

## Results

### Classifications of CWDB

By analyzing the literature on the morphology of CWDB, L-form, and mycoplasmas, we found that some bacterial forms in our samples matched the descriptions found in those studies. Based on these descriptions, we classified various types of CWD bacteria into the following forms and tested the prevalence of each form in our study population:

The protoplastic form varies from tiny motile granules (< 1 μm) to large spheric formations (up to 30 μm). The spheric form is the most frequent and can contain vesicles or granules (motile or not) (Fig. 1 a, Figs. 6 a,b,e-h, Movie S1, Movie S2). Due to the plasticity of the cellular membrane, some forms appear elongated or ovoid. Fig. 1b shows a high prevalence of spheric forms. Analyzing videos of samples we were able to capture the transformation of a spirochete-like in a protoplastic forms (Figs. 3 a-c, Movie S8). We also used atomic force microscope (AFM) to examine CWDB at higher magnification, Figure 3d shows spirochete with spheric form of CWDB attached. Staining with Hoechst (Figs. 3 e-j) indicated an inhomogeneous distribution of DNA as genetic material within larger L-form cells.

**Figure 1.**
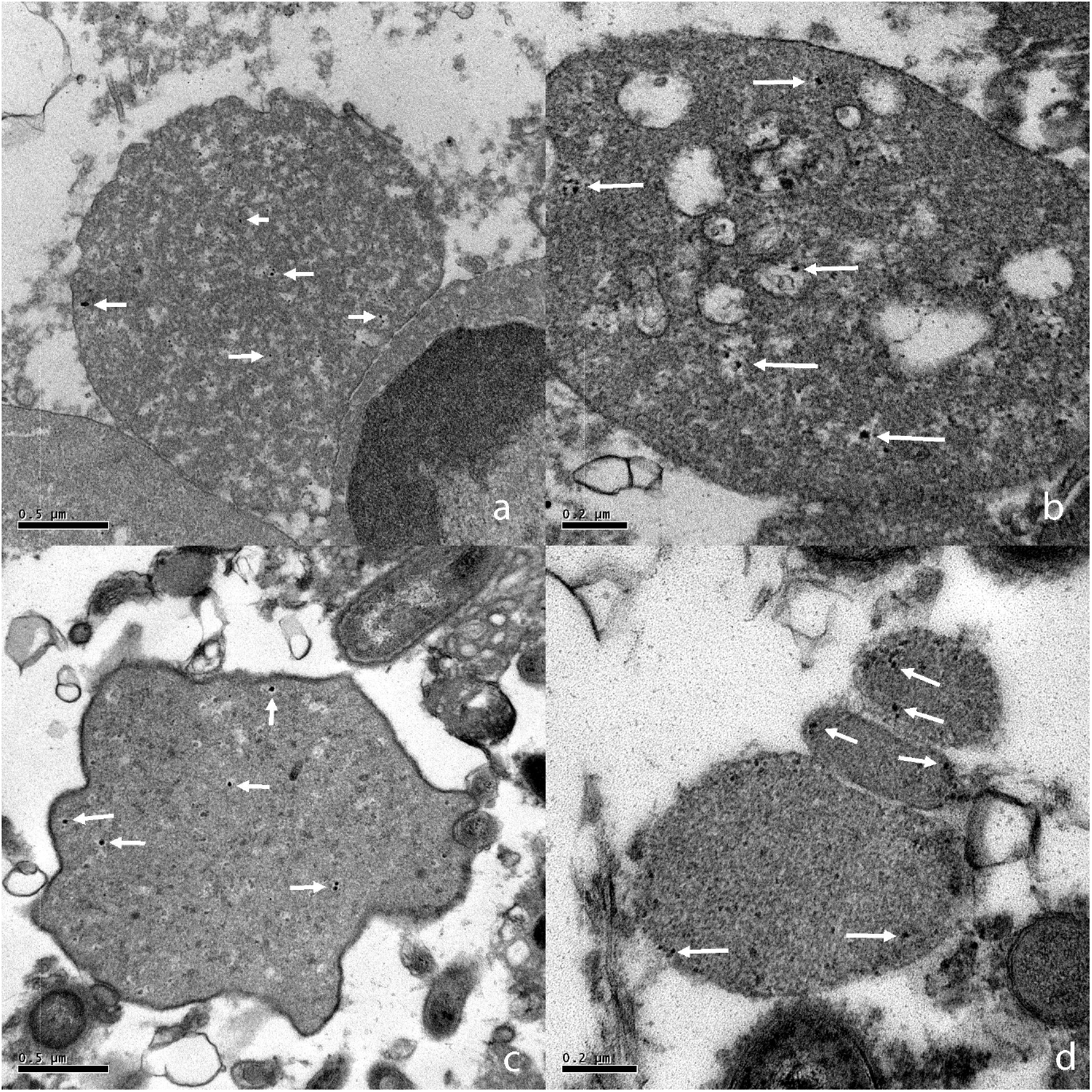
Protoplastic, everted, and filament forms of CWDB. **(a)** Low prevalence of the protoplastic form, arrows indicate forms with motile granule inside. **(b)** High prevalence of protoplastic tiny motile granules (< 1 μm) and large spheric formations (> 30 μm, see arrows). **(c)** Filament-form long, tiny branched formation connected with some protoplastic forms. **(d)** Everted form with tubular and bubble formations emerging from plaque aggregate. Light microscope in phase contrast. Bar = 10 μm.

Filament forms have the shape of long, tiny, non-motile formations (0.5-1 μm diameter, more than 100 μm long). Some of these are branched, connected with granulocytes or protoplastic forms (Fig. 1 c).

Everted forms originate from the surfaces of bacterial aggregates and can be recognized by their long motile tubular or bubble form. Their size ranges from small elongated bubbles (5 μm) to long tubular formations (5-10 μm diameter, more than 100 μm long). Some of these forms show motile granules inside the external membrane (Fig. 1 d, Movie S3).

The intracellular forms may be present in granulocytes, keratinocytes, and erythrocytes (Fig. 2). CWDB present in leukocytes have the shape of tiny and highly motile granules. Invaded granulocytes have the tendency to die by necrosis and to release CWDB into the surrounding space (Fig. 2 b, Movie S4). CWDB present in keratinocytes have the shape of tiny and highly motile granules (Fig. 2 a, Movie S6). They are located in cytoplasmic space and can sometimes be found inside nuclear space (Fig. 2d-e, Movie S5). CWDB were also seen attached to the surface of erythrocytes in the form of motile protrusions that have the shape of long pearl-like chains extending up to 30 μm from the cell (Fig. 2 c, Movie S7). We also included images and videos of samples obtained from periodontally healthy individuals (Figs. 4 a-b) and chronic periodontitis (Figs. 4 c-d, Figs. 7 a-d, Movie S9) which do not contain CWDB bacteria but show only normal bacterial morphology. No differences were seen between biofilms dispersed in saline solution and in osmoprotective media.

**Figure 2.**
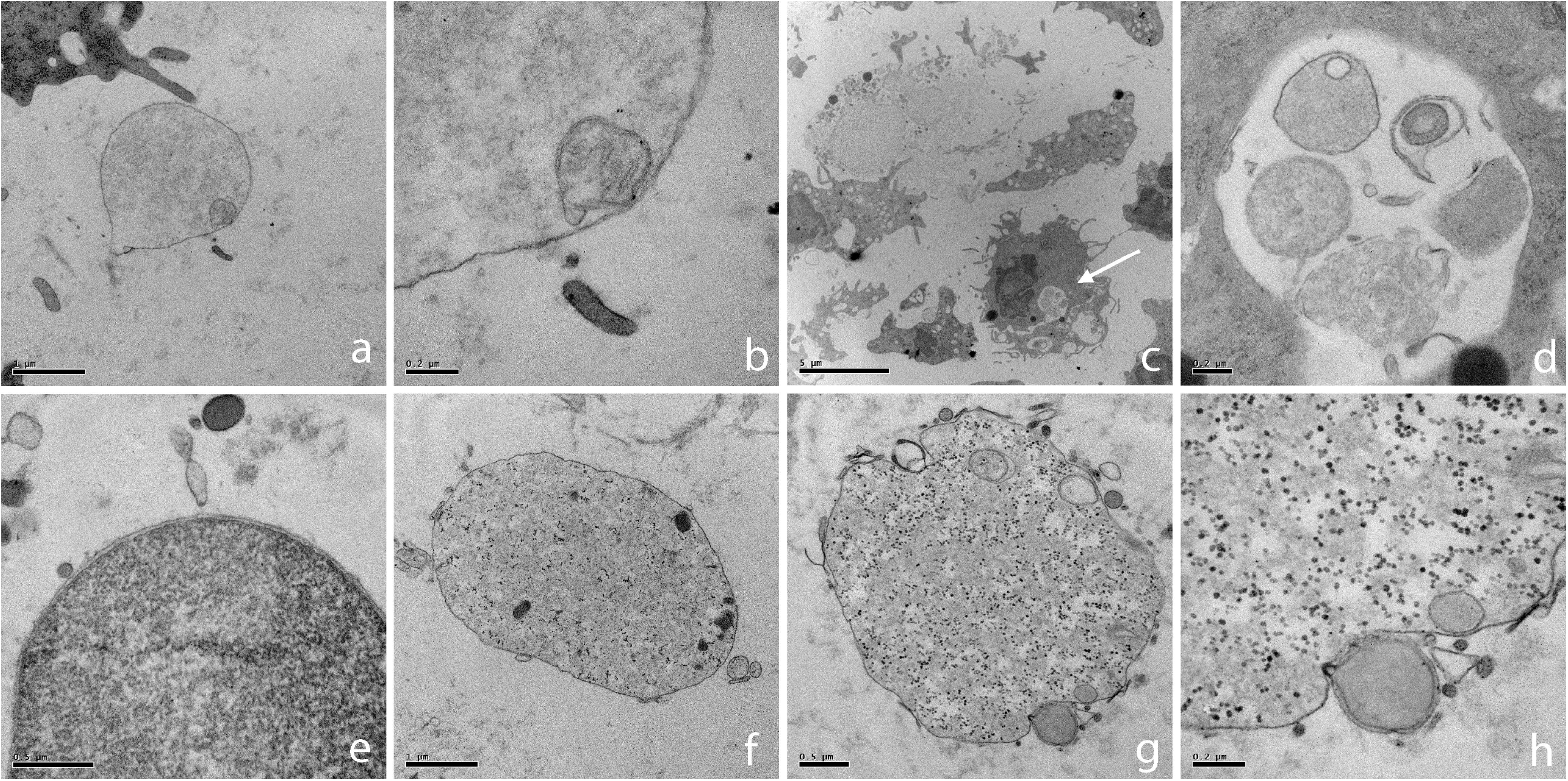
Intracellular form of CWDB. **(a)** CWDB inside cytoplasmic space of a keratinocyte. Arrows indicate motile CWDB. **(b)** Time-lapse photography of a granulocyte invaded by CWDB, releasing tiny forms of bacteria into the surrounding space. **(c)** CWDB present in erythrocytes. Arrows indicate motile chain of CWDB. **(d)** CWDB inside nuclear space of a keratinocyte. Arrows indicate motile CWDB. Light microscope in phase contrast. **(e)** CWDB inside cytoplasmic space of a keratinocyte Hoechst immunofluorescence staining. Bar = 10 μm.

**Figure 3.**
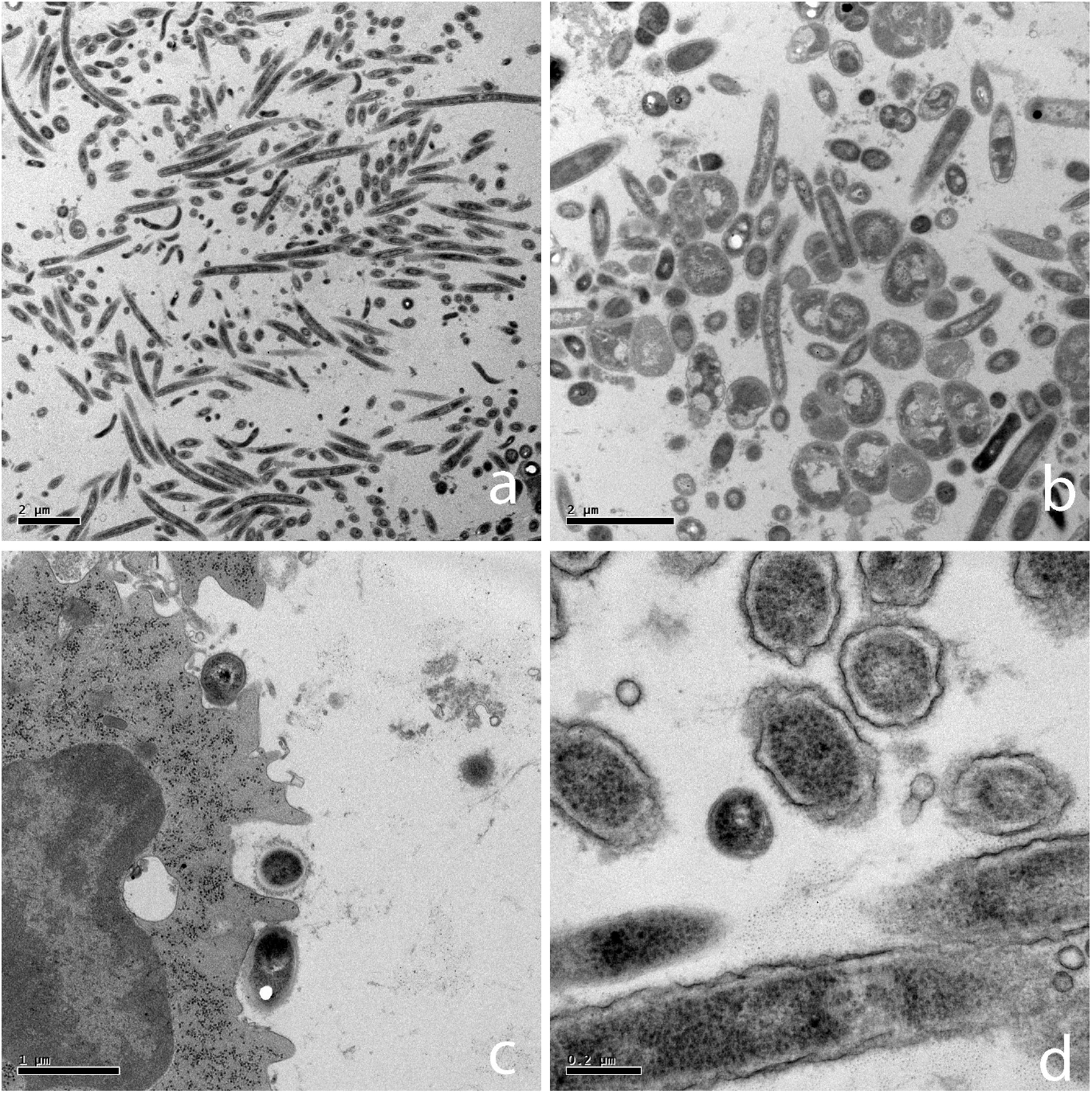
**(a-c)**. Time-lapse photography of the transformation of a spirochete-like bacteria in a spheric form of CWDB Light microscope in phase contrast.. **(d)** Spirochete with spheric form of CWDB attached. Atomic force microscopy. Image height is 0-140 nm. **(e-J)** Hoechst immunofluorescence staining indicating small vesicles and inhomogeneous distribution of DNA. Bar = 10 μm.

**Figure 4.**
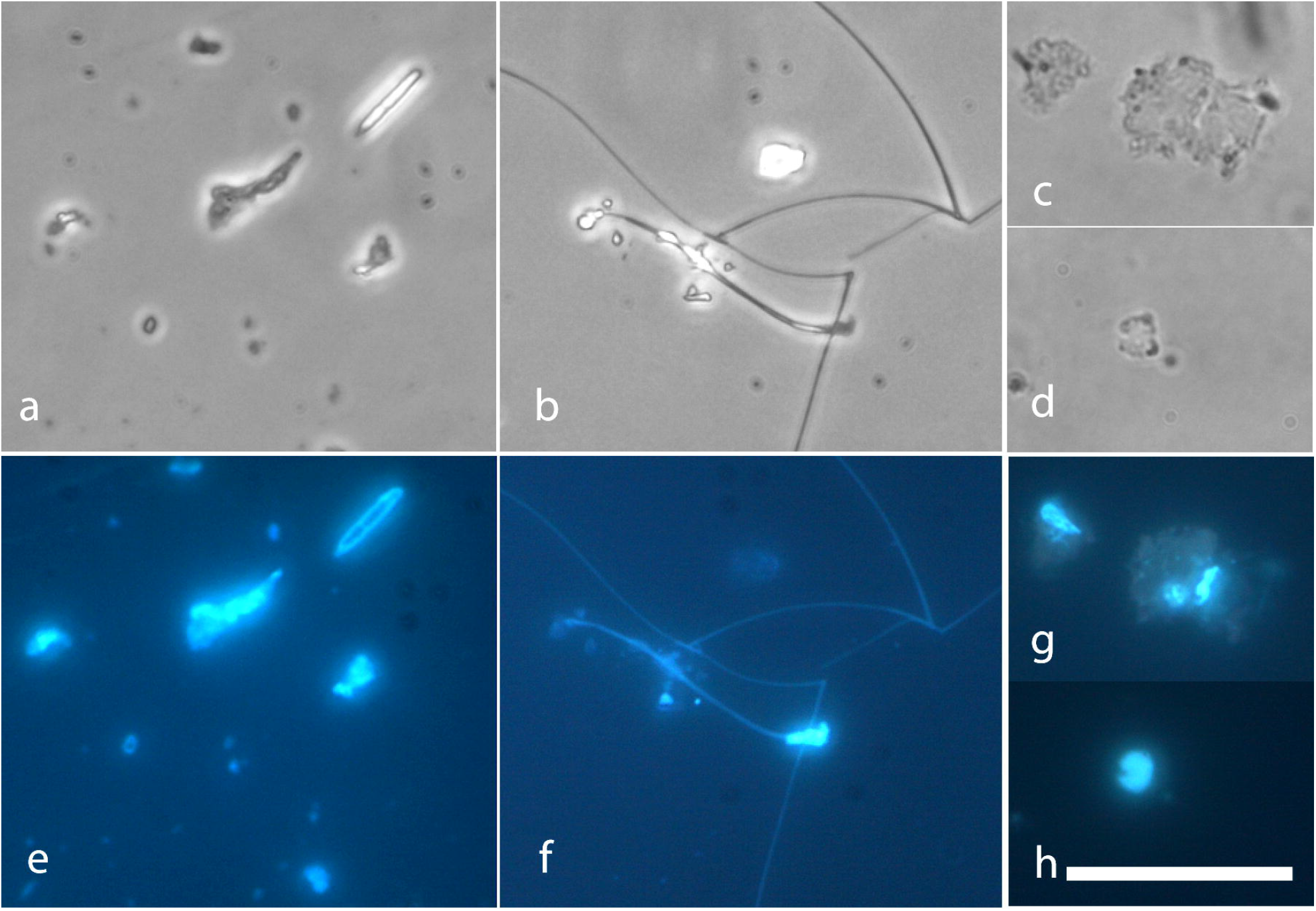
Control images with classical bacterial forms. Periodontally healthy individuals **(a)** corncob formation, **(b)** “test-tube brush” formation, **(c)** coccoid and straight rod bacteria. Chronic periodontitis individuals **(d)** all major morphological categories are represented (cocci, straight rods, filament, spirochete), a granulocyte is also present in the upper half. Bar = 10 μm.

In samples processed for electron microscopy, CWDB were seen only in aggressive periodontitis individuals (Figs. 6 a-h) while in chronic periodontitis individuals only walled bacteria were found (Figs. 7 a-d). A variety of forms were found in each aggressive periodontitis sample. Range sizes vary from 0.2 μm to 10 μm. CWDB can be recognized by the presence of the plasma membrane bi-layer and the absence of the cell wall, as shown in Figure 6e. In some cases these organisms presented a light granular cytoplasm with small vesicles inside, possibly representing reproductive units (Fig 6 a,b,g,h). The presence of small ribosome-like structure was also observed (Fig 6 f-h). CWDB were found inside granulocytes, and a representative image of many pleomorphic small forms inside a vacuole is shown in Figures 6 c,d. Larger CWDB presented small dark and light vesicles inside the cytoplasm and some could be seen outside the membrane layer (Fig 6 g,h).

In samples processed for TEM with electron microscopy in situ hybridization, presence of bacterial DNA was seen in CWDB from aggressive periodontitis individuals (Figs 8 a-d). EDX microanalysis was also applied to confirm presence of gold particles (Data not shown)

**Figure 5.**
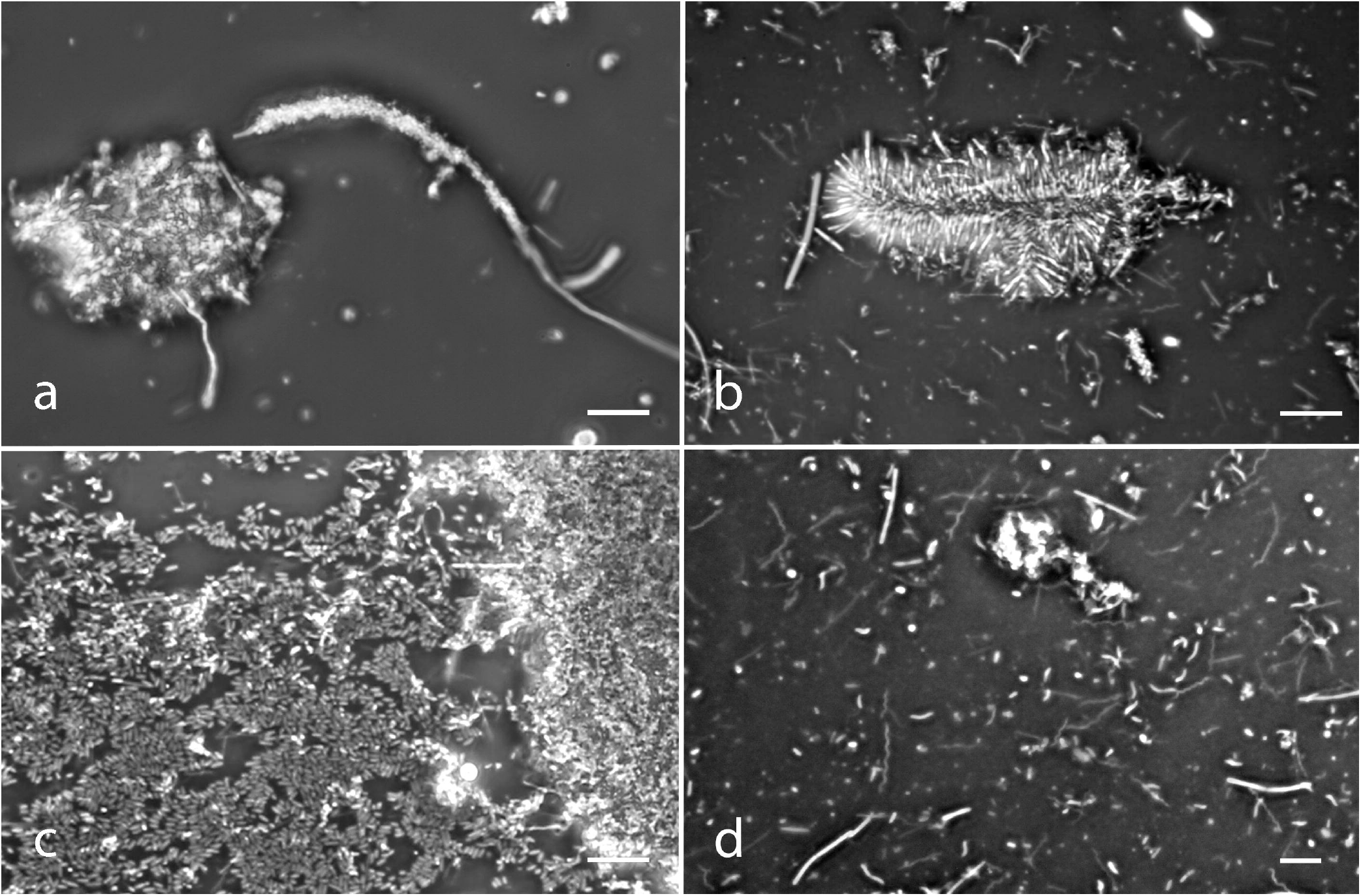
CWDB after culture and filtration with 0.45 μm filter. **(a)** Protoplastic form. (b) Filament form. **(c**,**d)** Damaged protoplastic form. **(e-h)** Hoechst immunofluorescence staining of images above. Bar = 5 μm

**Figure 6.**
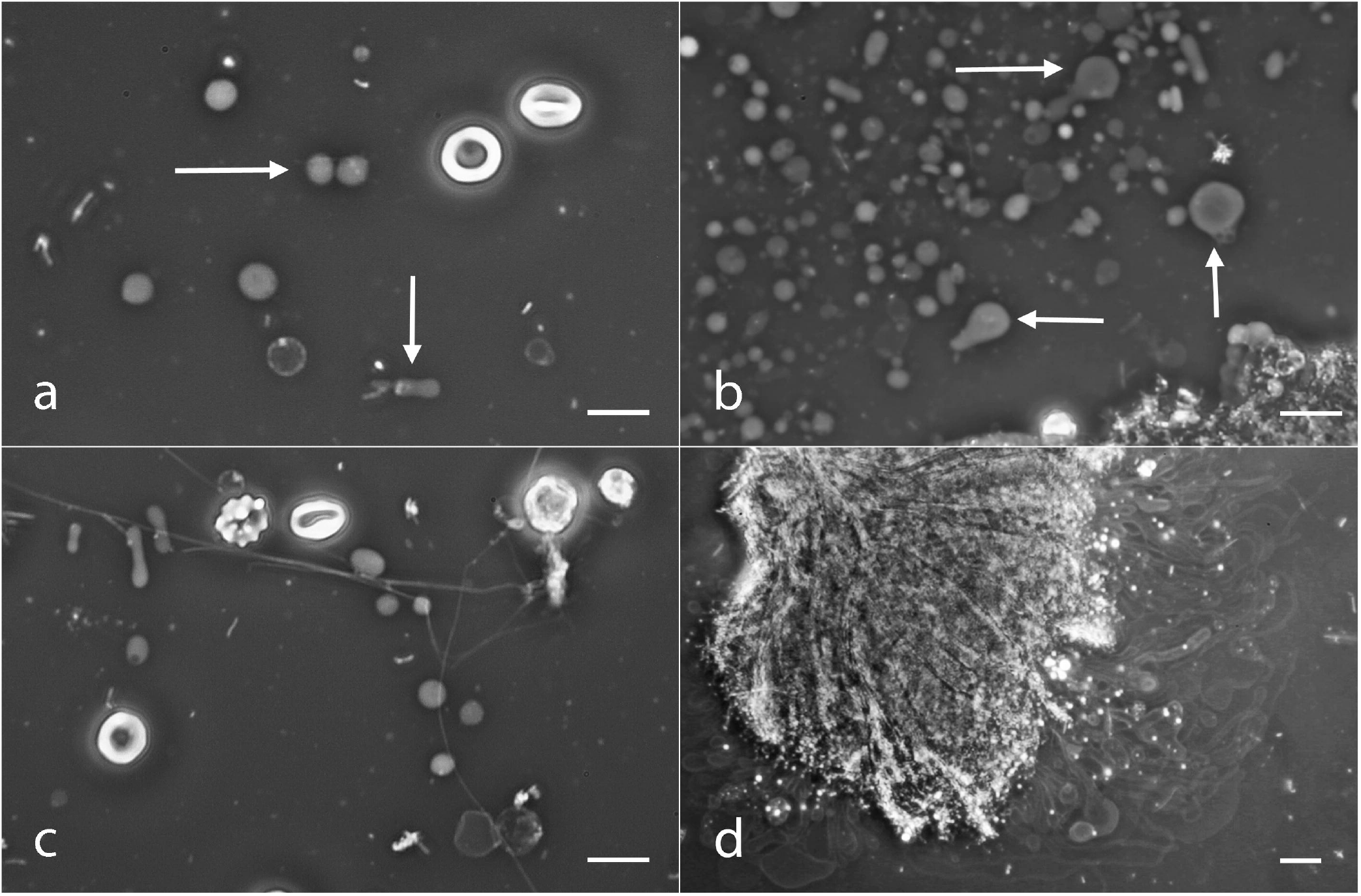
TEM Images of CWDB from Aggressive Periodontitis Individuals. (a) CWDB with light granular cytoplasm and a small vesicle inside. 20000x. (b) Vesicle at higher magnification, double layer membrane is easy visible. 80000x. (c) A granulocyte containing a CWDB inside a vacuole (arrow); mag. 5000x. (d) A higher magnification of the same vacuole shown in c. CWDB with lighter or darker cytoplasm can be observed inside the vacuole. 60000x. (e) CWDB with dark granular cytoplasm and a well conserved plasma membrane. Light and dark small vesicles are present outside cell membrane. 50000x. (f) Large CWDB (up to 5 μm) with ribosome-like granules and dark inclusions. 20000x. (g) CWDB with ribosome-like granules. Light and dark small vesicles are present inside and outside cell membrane. 30000x. (h) Magnification of previous image; 80000x.

**Figure 7.**
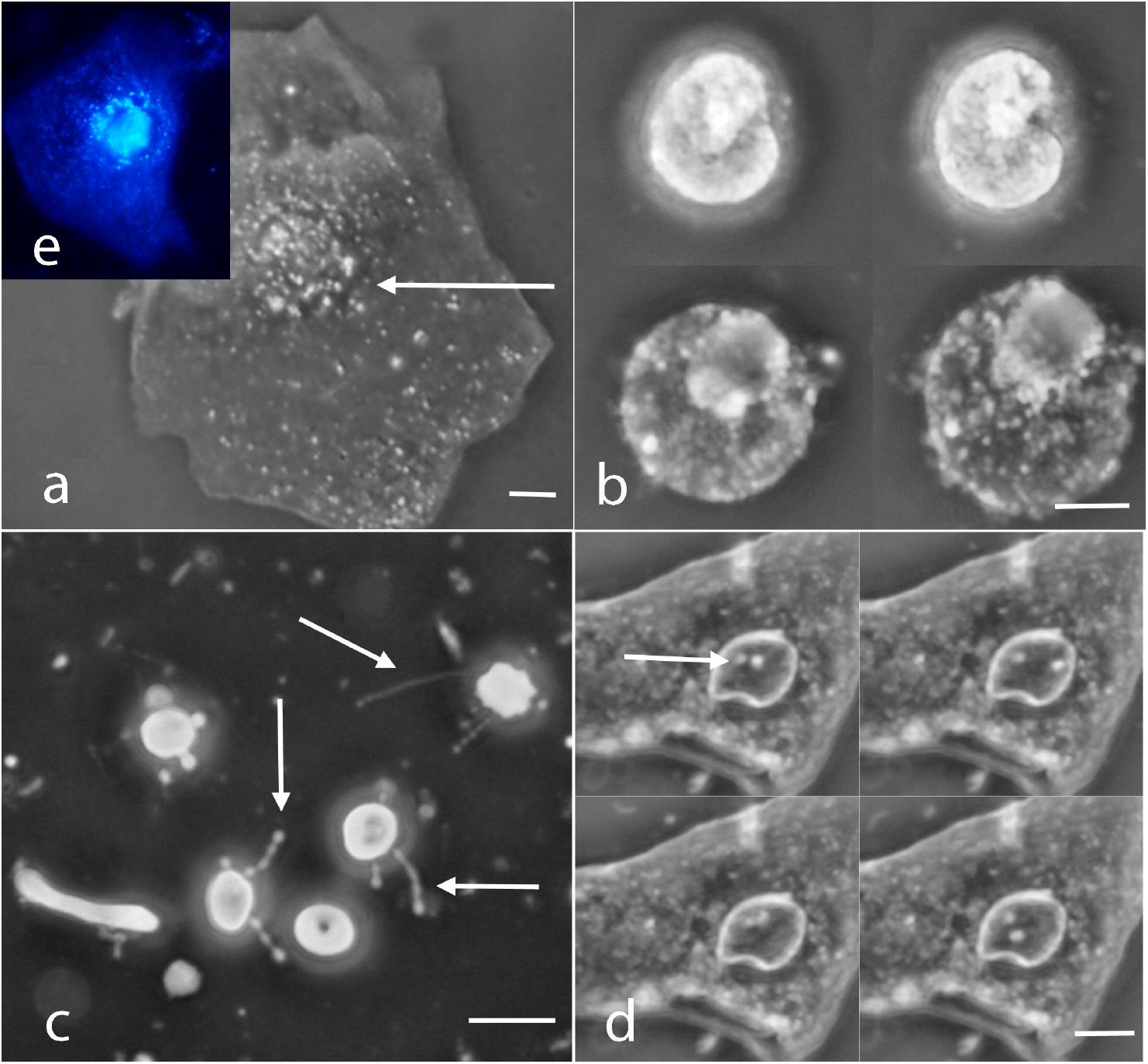
TEM Images of classical walled bacteria from chronic Periodontitis Individuals. (a) Coccoid, Fusiform, and few Spirocheta bacteria with classical cell wall morphology. 6000x. (b) Coccoid and Fusiform walled bacteria; 10000x. (c) Leucocyte with various walled bacteria attached to his plasma membrane; 20000x. (d) Gram-bacteria 80000x

**Figure 8.**
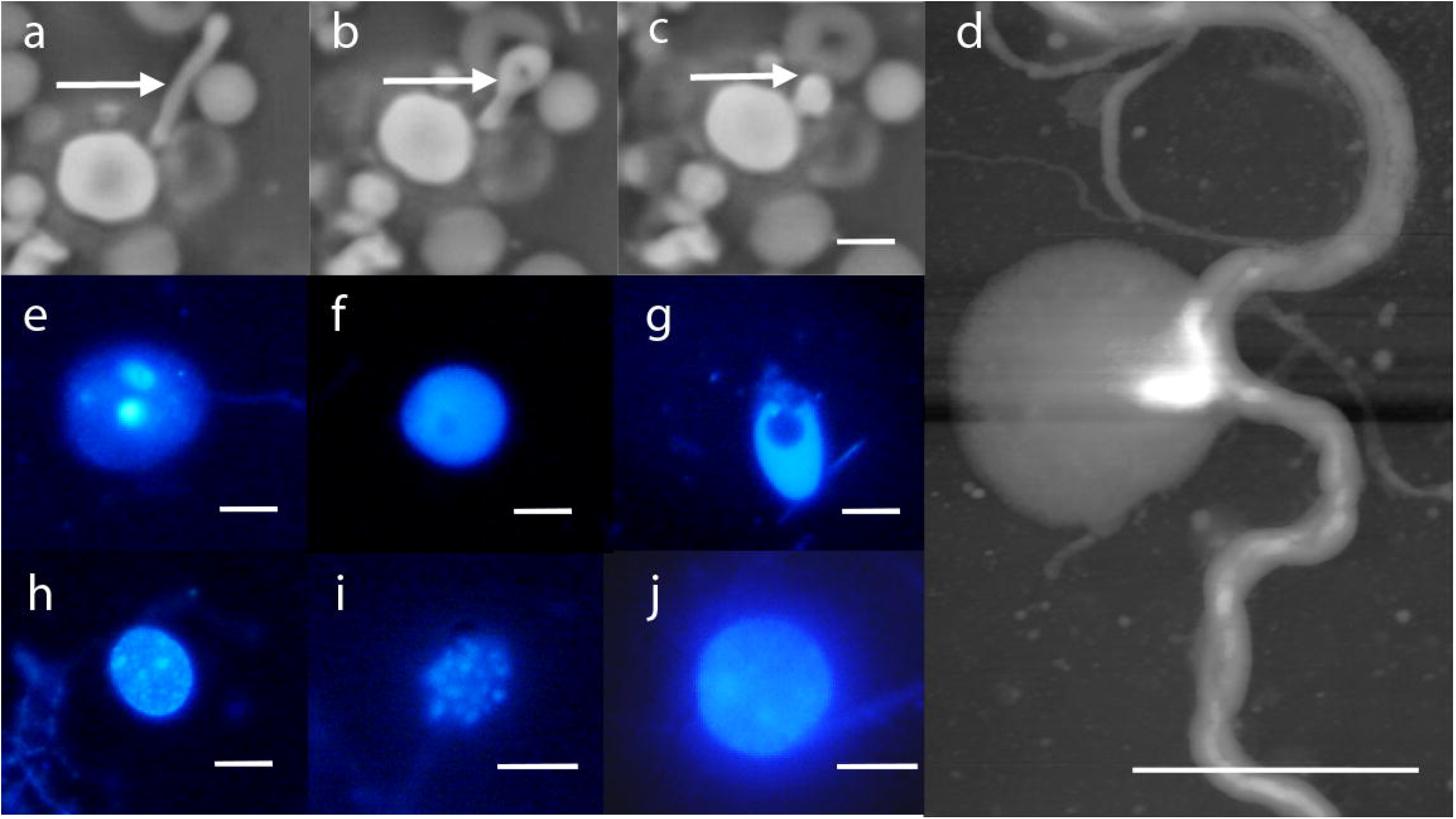
**(a-c)** TEM In Situ Hybridization Images (EUB 338 for 16S rRNA of most bacteria) of CWDB from Aggressive Periodontitis Individuals (arrows indicate gold particles). **(a)** CWDB with several gold particles inside cytoplasmic space adjacent to the membrane of leucocyte 50000X. **(b)** CWDB with vacuoles inside cytoplasm 80000x. **(c)** CWDB with a well conserved plasma membrane surrounded by walled bacteria 50000x. **(d)** Three small CWDB 80000x.

When we analyzed biofilm culture samples, 2 from periodontally healthy individuals (G0), 2 from chronic periodontitis individuals (G0 and G1L) and 3 from aggressive periodontitis individuals (two G1L and one G1H), only in samples classified G1L and G1H we observed CWDB forms after filtration through 0,45 μm filters to exclude walled bacteria (Fig. 5). In one filtered G1H sample we also detected motile CWDB (Movie S10)

### Statistical Correlation between CWDB and Individuals with Chronic or Aggressive Periodontal Disease and with Healthy Individuals

A sample of 89 individuals was evaluated (30 periodontally healthy, 36 with chronic periodontitis, and 23 with aggressive periodontitis). Fig. 9 shows the number of patients presenting with CWDB.

**Figure 9.**
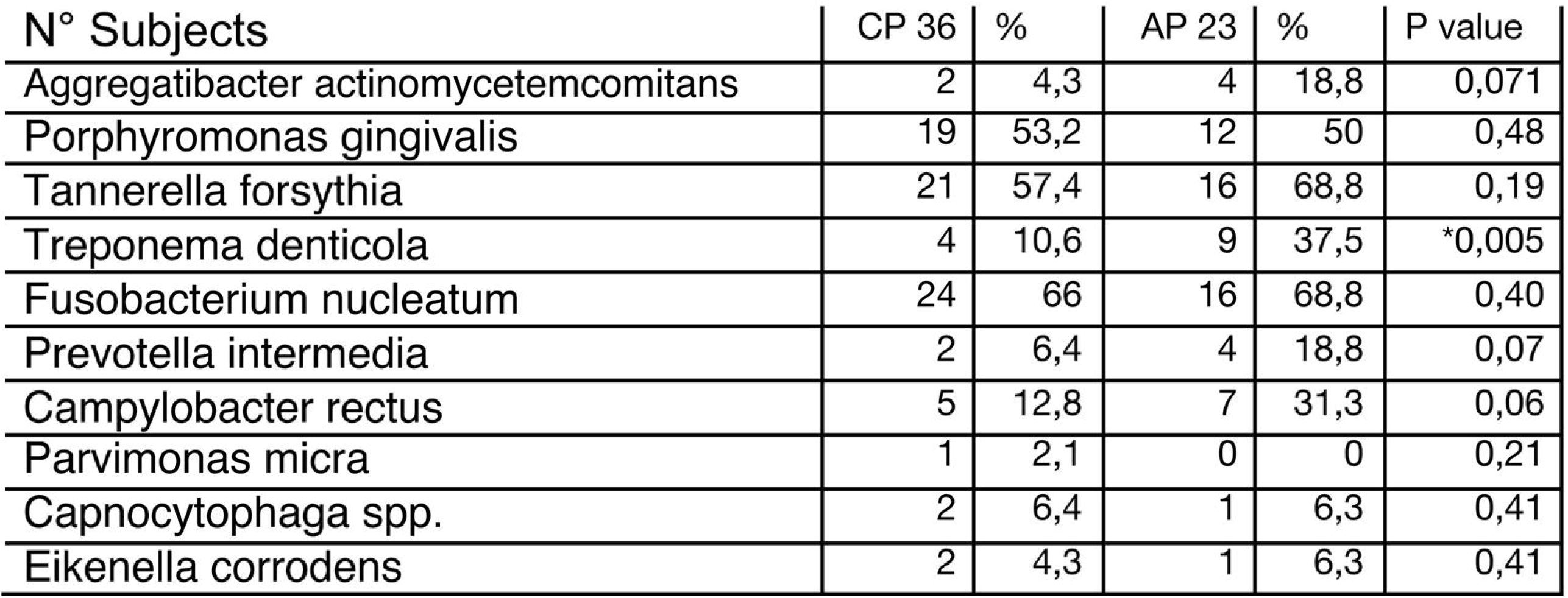
Association between CWDB and Periodontal Condition. G0: absence of CWDB or the presence of only protoplastic form without granules G1L: 2 kinds of CWDB were present or CWDB prevalence > 5%; G1H: 3 or more kinds of CWDB were present, or high prevalence of CWDB were found (> 70%). (see text for details) Periodontally Healthy (HP, n = 30), G0 = 29 (96.7%), G1L = 1 (3.3%), G1H = 0 (0%). Chronic periodontitis (CP, n = 36), G0 = 25 (69.4%), G1L = 10 (27.8%), G1H = 1 (2.8%). Aggressive periodontitis (AP, n = 23), G0 = 0 (0%), G1L = 12 (52.2%), G1H = 11 (47.8%).

The results, listed below, were established after descriptive statistical evaluations were performed.

The three groups showed the following values of CWDB: HP, G1 = 1 (3.3%); CP, G1 = 11 (30.6%); and AP, G1 = 23 (100%). The differences among the groups were statistically significant. Marked differences were found between HP and AP (chi-square = 49.10, *p* <.0001), between CP and AP (chi-square = 27.52, *p* < .0001), and between HP and CP (chi-square = 8.152, *p* = .00022). If we divide the G1 score in G1L and G1H, we have the highest G1H count in the AP group (G1L = 12, G1H = 11), while in CP only one sample presented this score (G1L = 10, G1H = 1).

To test the significance among the four kinds of CWDB we classified, we assessed the presence of each form in CP and AP groups positive for CWDB. We found no statistically significant correlation in the protoplastic, filament, and everted forms between the groups. Only intracellular forms showed a statistically significant correlation with the AP group (chi-square, 3.387; *p* = .032).

### PCR

To verify if CWDB are associated with known periodontal pathogenic bacteria, we assessed the prevalence of periodontopathic bacteria by means of polymerase chain-reaction (PCR). We found no clear, statistically significant, correlation among periodontal pathogens tested (except *T. denticola, p* = .005) that allowed individuals with chronic periodontitis to be distinguished from those with aggressive periodontitis (Fig. 10).

**Figure 10.**
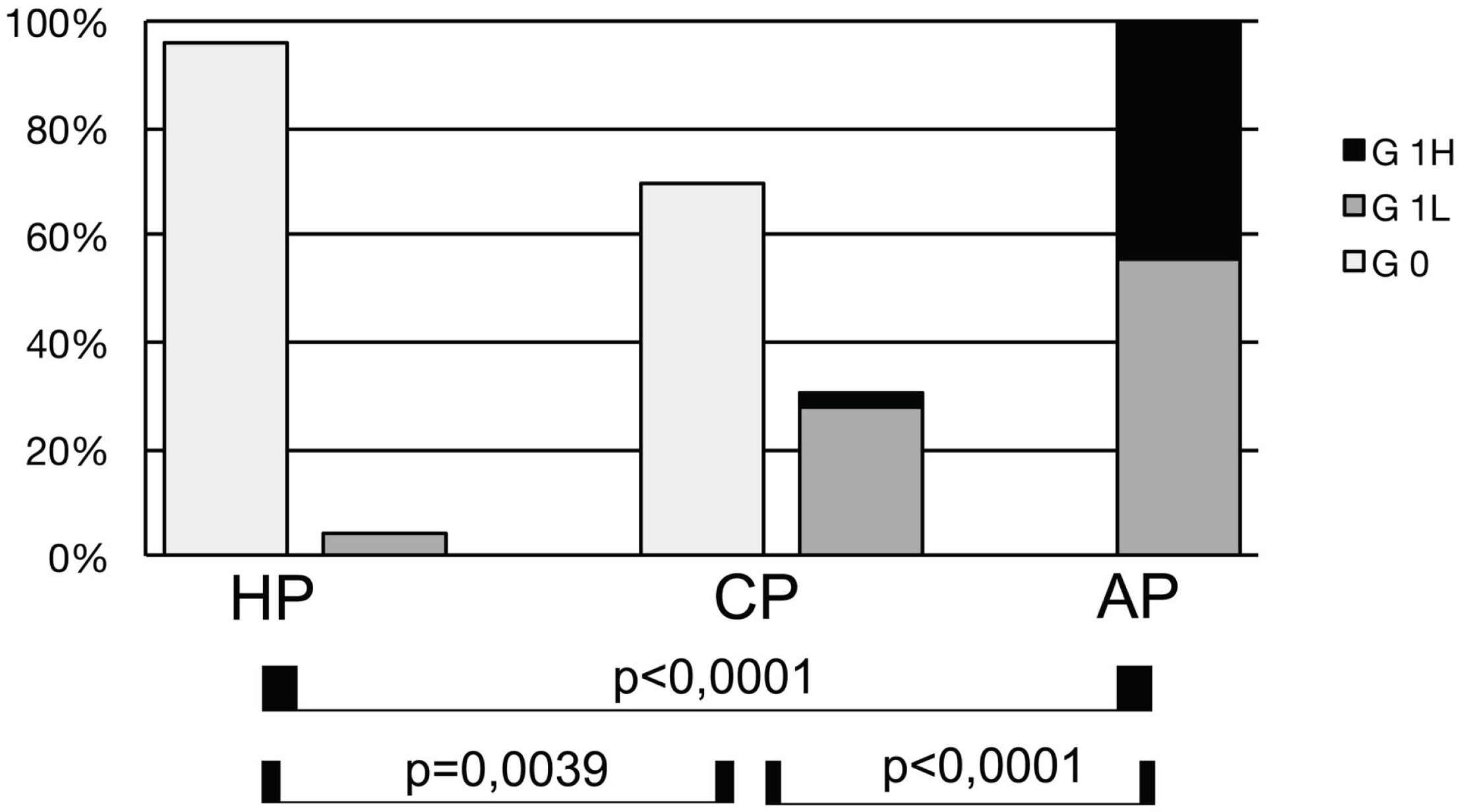
PCR of Major Periodontopathic Bacteria Analyzed in Chronic (CP) and Aggressive (AP) Periodontitis (P = significance of chi-square analysis, * statistically significant)

## Discussion

Literature regarding CWDB in the oral cavity is not up to date and is based on unclear classifications, with terms such as PPLO, mycoplasma, spheroplast, protoplast, and L-form often describing the same bacterial form. The majority of studies classifying and analyzing CWDB used sophisticated culture techniques to select and maintain this bacterial state. To our knowledge, this is the first study linking CWDB to periodontal disease.

To overcome the difficulty and bias of the bacterial culture technique, we developed a simple method to verify morphology on fresh biofilm as well as the presence and prevalence of CWDB in periodontal disease according to the current classification, in which chronic and aggressive periodontitis are separate diseases. Since we know from the literature that a multitude of, if not all, bacteria can assume the CWD form, a molecular study to characterize those forms of bacteria in our sample was beyond the scope of this research.

The results demonstrated statistically significant results confirming a considerable association between CWDB and aggressive periodontitis, with a low association in periodontally healthy individuals and in those with chronic periodontitis.

Another important observation was the presence of varieties related to the kind of periodontal disease: everted forms, filament forms, areas containing more than 80% of CWDB, erythrocyte associated forms and endonuclear invasion were consistently related to aggressive periodontitis (data not shown).

Our analyses of the relation between CWDB and healthy individuals and those with chronic and aggressive periodontitis showed that only aggressive periodontitis was highly associated with CWDB grade 1 values (100%, 23/23), while the associations in individuals with chronic periodontitis was low (30.6%, 11/36) and periodontally healthy individuals (4%, 1/30) was very low.

These results could be connected to differences in terms of progression of periodontal destruction and severity of aggressive and chronic disease.

This was not surprising, because the transformation in CWDB can alter bacterial gene expression and can amplify virulence factors such as: horizontal gene transfer, capacity to penetrate host cells, stealthy entrance into the immune system. We now know that CWDB requires a combination of two kinds of mutations, one work by generating excess amounts of cell membrane and another work by stabilizing the proliferating CWDB and preventing them from lysing. Oxidative damage prevents growth of CWDB and reduction of ROS levels or anaerobic conditions promotes their growth (Kawai at al 2015).

TEM images obtained from aggressive periodontitis samples showed that the variety of forms that CWDB take are similar to images seen, at lower magnification, with phase contrast microscope and to the ultrastructure of Mycoplasma and L-forms of bacteria [19-20].

Another interesting observation was the inhomogeneous distribution of DNA in the protoplastic forms, as tested with Hoechst staining, that was comparable with images obtained in *Listeria monocytogenes* cultures indicating intracellular vesicles as reproduction elements [30].

To test whether CWDB seen were true bacteria electron microscopy In Situ Hybridization with the EUB 338 probe, specific for general Eubacteria, on 9 samples (3 for each clinical groups analyzed), only samples from aggressive periodontitis individuals shows CWDB. Which mechanisms induce the loss of cell walls in these bacteria? Some studies suggest that a component in the membrane of the L-form, distinct from cell wall components such as teichoic acid or lipopolysaccharide, possesses the capability to stimulate TNF-α production by macrophages [31-35]. The mechanisms of macrophage activation by the membranes of mycoplasmas, were shown to increase TNF-α production [33].

Another study has demonstrated that a ‘hyperresponsive’ phenotype in individuals with localized aggressive periodontitis, upon activation by common periodontal pathogens, could contribute to exacerbated inflammatory responses and eventual rapid host tissue destruction [36].

Another study [37] indicated differential immunoregulatory mechanisms in AP and CP, suggesting the presence of different cellular activation in the two different clinical forms. These mechanisms could be related to distinct bacteria or antigens present during disease. Our study results add another small piece to the multifactorial relationship between periodontal biofilm and host response, suggesting that CWDB are associated mainly with aggressive periodontitis. Further studies are needed for molecular characterization of these bacteria and clarification of whether CWDB are an effect of a specific host response or are capable of altered immune response.

Ultimately we believe that knowledge of the role of CWDB in periodontal disease could improve or help the diagnosis and development of new therapeutic protocols for these patients.

## Data Availability

Individual data (PD, CAL, PI, BOP, etc.) for all patients are presented in the online Supporting Information (Table S1)

## Acknowledgments

This research received no specific grant from any funding agency in the public, commercial, or not-for-profit sector. The authors declare that they have no conflicts of interest.

## References

[1] Errington J. L-form bacteria, cell walls and the origins of life. Open Biol. 2013; 3:120143.

[2] Chen, T., Yu, W-Han, Izard, J., Baranova, O.V., Lakshmanan, A., Dewhirst, F.E. (2010) The Human Oral Microbiome Database: a web accessible resource for investigating oral microbe taxonomic and genomic information. Database, Vol. 2010, Article ID baq013, doi:10.1093/database/baq013.

[3] Domingue GJ Sr, Woody HB. Bacterial persistence and expression of disease. Clin Microbiol Rev. 1997; 10:320–344.

[4] Onwuamaegbu ME, Belcher RA, Soare C. Cell wall-deficient bacteria as a cause of infections: a review of the clinical significance. J Int Med Res. 2005; 33:1–20.

[5] Razin S, Yogev D, Naot Y. Molecular biology and pathogenicity of mycoplasmas. Microbiol Mol Biol Rev. 1998; 62:1094–1156.

[6] Razin S, Michmann J, Shimshoni Z. The occurrence of mycoplasma (pleuropneumonia-like organisms, PPLO) in the oral cavity of dentulous and edentulous subjects. J Dent Res. 1964; 43:402–405.

[7] Watanabe T, Matsuura M, Seto K. Enumeration, isolation, and species identification of mycoplasmas in saliva sampled from the normal and pathological human oral cavity and antibody response to an oral mycoplasma (Mycoplasma salivarium). J Clin Microbiol. 1986; 23:1034–1038.

[8] Kwek HSN, Wilson M, Newman HN. Mycoplasma in relation to gingivitis and periodontitis. J Clin Periodontol. 1990; 17:119–122.

[9] Newman MG, Saglie R, Carranza F, Kaufman AK. Mycoplasma in periodontal disease: isolation in juvenile Periodontitis. J Periodontol. 1984; 55:574

[10] Kleinberger E. The natural occurrence of pleuropneumonia-like organisms in apparent symbiosis with Streptobacillus moniliformis and other bacteria. J Pathol Bacteriol. 1935; 40:93–105.

[11] Allan EJ, Hoischen C, Gumpert J. Bacterial L-forms. Adv Appl Microbiol. 2009; 68:1–39.

[12] Mercier R, Kawai Y, Errington J. Excess membrane synthesis drives a primitive mode of cell proliferation. Cell. 2013 28;152(5):997–1007. doi:10.1016/j.cell.2013.01.043.

[13] Cambre A, Zimmermann M, Sauer U, Vivijs B, Cenens W, Michiels WC, Aertsen A, Loessner JM, Noben J, Ayala, AJ, Lavigne R, Briers Y. Metabolite profiling and peptidoglycan analysis of transient cell wall-deficient bacteria in a new Escherichia coli model system. Environ Microbiol. 2014; http://dx.doi.org/10.1111/1462-2920.12594

[14] Markova N, Slavchev G, Michailova L, Jourdanova M. Survival of Escherichia coli under lethal heat stress by L-form conversion. Int J Biolog Sci. 2010; 6:303–315.

[15] Kagan GY, Mikhailova VS. Isolation of L-forms of streptococci from blood of patients with rheumatism and endocarditis. J Hyg Epidemiol Microbiol Immunol. 1963; 13:327–343.

[16] Gordon SL, Greer RB, Craig CP. Recurrent osteomyelitis: report of four cases culturing L form variants of staphylococci. J Bone Joint Surg Am. 1971; 53:1150–1156.

[17] MacDonald AB. Concurrent neocortical borreliosis and Alzheimer’s disease: demonstration of a spirochetal cyst form. Ann NY Acad Sci. 1988; 539:468–470.

[18] Macomber PB. Cancer and cell wall deficient bacteria. Med Hypotheses. 1990; 32:1–9.

[19] Anderson DR, Barile M. Ultrastructure of Mycoplasma hominis. J Bacteriol. 1965; 90:180–192

[20] Dienes L, Bullivant S. Morphology and reproductive processes of the L forms of bacteria. II. Comparative study of L forms and Mycoplasma with the electron microscope. J Bacteriol. 1968; 95(2):672–687.

[21] Mombelli A, Casagni F, Madianos PN. Can presence or absence of periodontal pathogens distinguish between subjects with chronic and aggressive periodontitis? A systematic review. J Clin Periodontol. 2002; 3:10–21.

[22] Armitage GC. Development of a classification system for periodontal diseases and conditions. Ann Periodontol. 1999; 4:1–6.

[23] Germano F, Bramanti E, Arcuri C, Cecchetti F, Cicciù M. Atomic force microscopy of bacteria from periodontal subgingival biofilm: preliminary study results. Eur J Dent. 2013; 7:152–158.

[24] Armitage GC, Cullinan MP. Comparison of the clinical features of chronic and aggressive periodontitis. Periodontol 2000. 2010; 53:12–27

[25] Demmer RT, Papapanou PN. Epidemiologic patterns of chronic and aggressive periodontitis. Periodontol 2000. 2010; 53:28–44

[26] Eick S, Pfister W. Comparison of microbial cultivation and a commercial PCR based method for detection of periodontopathogenic species in subgingival plaque samples. J Clin Periodontol. 2002; 29: 638–644.

[27] Scimeca M, Giannini E, Antonacci C, Pistolese CA, Spagnoli LG, Bonanno E. Microcalcifications in breast cancer: an active phenomenon mediated by epithelial cells with mesenchymal characteristics. BMC Cancer. 2014; Apr 23;14:286. doi:10.1186/1471-2407-14-286.

[28] Reynolds ES. The use of lead citrate at high pH as an electron opaque stain based on metal chelation. J Cell Biol. 1963; 17:208–212

[29] Mattman LH. Stealth pathogens.. 2001; 3rd ed. Boca Raton, FL: CRC Press.

[30] Briers Y, Staubli T, Schmid MC, Wagner M, Schuppler M, Loessner MJ. Intracellular Vesicles as Reproduction Elements in Cell Wall-Deficient L-Form Bacteria. PLoS ONE. 2012; 7(6): e38514. doi:10.1371/journal.pone.0038514

[31] Gallily R, Sher T, Ben-Av P, Loewenstein J. Tumor necrosis factor as a mediator of Mycoplasma orale-induced tumor cell lysis by macrophages. Cell Immunol. 1989; 121:146–153.

[32] Arai S, Furukawa M, Munakata T, Kuwano K, Inoue H, Miyazaki T. Enhancement of cytotoxicity of active macrophages by mycoplasma: role of mycoplasma-associated induction of tumor necrosis factor-α (TNF-α) in macrophages. Microbiol Immunol. 1990; 34:231–243.

[33] Sher T, Rottem S, Gallily R. Mycoplasma capricolum membranes induce tumor necrosis factor α by a mechanism different from that of lipopolysaccharide. Cancer Immunol Immunother. 1990; 31:86–92.

[34] Sugama K, Kuwano K, Furukawa M, Himeno Y, Satoh T, Arai S. Mycoplasma induce transcription and production of tumor necrosis factor in a monocytic cell line, THP-1, by a protein kinase C-independent pathway. Infect Immun. 1990; 58:3564–3567.

[35] Kuwano K, Akashi A, Matsu-ura I, Nishimoto M, Arai S. Induction of macrophage-mediated production of tumor necrosis factor alpha by an L-form derived from Staphylococcus aureus. Infect Immun. 1993; 61:1700–1706.

[36] Shaddox LShaddox, L., Wiedey, J., Bimstein, E., Magnuson, I., Clare-Salzler, M., Aukhil, I., & Wallet, S. M. Hyper-responsive Phenotype in Localized Aggressive Periodontitis. Journal of Dental Research. 2010; 89(2), 143–148. doi:10.1177/0022034509353397

[37] Lima PM, Souza PE, Costa JE, Gomez RS, Gollob KJ, Dutra WO. Aggressive and chronic periodontitis correlate with distinct cellular sources of key immunoregulatory cytokines. J Periodontol. 2011; 82:86–95.

